# Factors associated with iron deficiency and how they can be used in blood donor selection processes

**DOI:** 10.1101/2022.02.01.22270122

**Authors:** Sofie Ekroos, Mikko Arvas, Johanna Castrén

## Abstract

**Background and objectives:** In an effort to improve donor health, we set out to investigate determinants other than blood donation that increase risk of ID in a healthy population and if they should be implemented into current blood donor selection processes.

**Methods:** We systematically searched the PubMed, Ovid Medline and Scopus for articles related to ID.

**Results:** Current evidence suggests that several determinants, including biological, environmental, lifestyle and socioeconomic factors increase the risk for ID.

**Conclusions:** Heavy menstruation, use of some medications and dietary factors could potentially be implemented in donor selection, however further study is needed.

## Introduction

The purpose of this systematic literature review was to investigate determinants for iron deficiency and/or iron deficiency anemia in the normal population, how they relate to blood donor selection processes and to which degree they could possibly be implemented in the donor selection process.

## Methods

Risk factors for both ID and IDA were chosen as the focus of the search. We applied an age restriction of 15-70 years and only included studies from countries with Human Development Index (HDI) ≥0.800 according to the 2020 data in order to produce a coherent data set (1).

### Systemic search

We selected three databases: PubMed, Ovid Medline and Scopus. The conducted searches and results are included in Table 1.

**Table 1:**
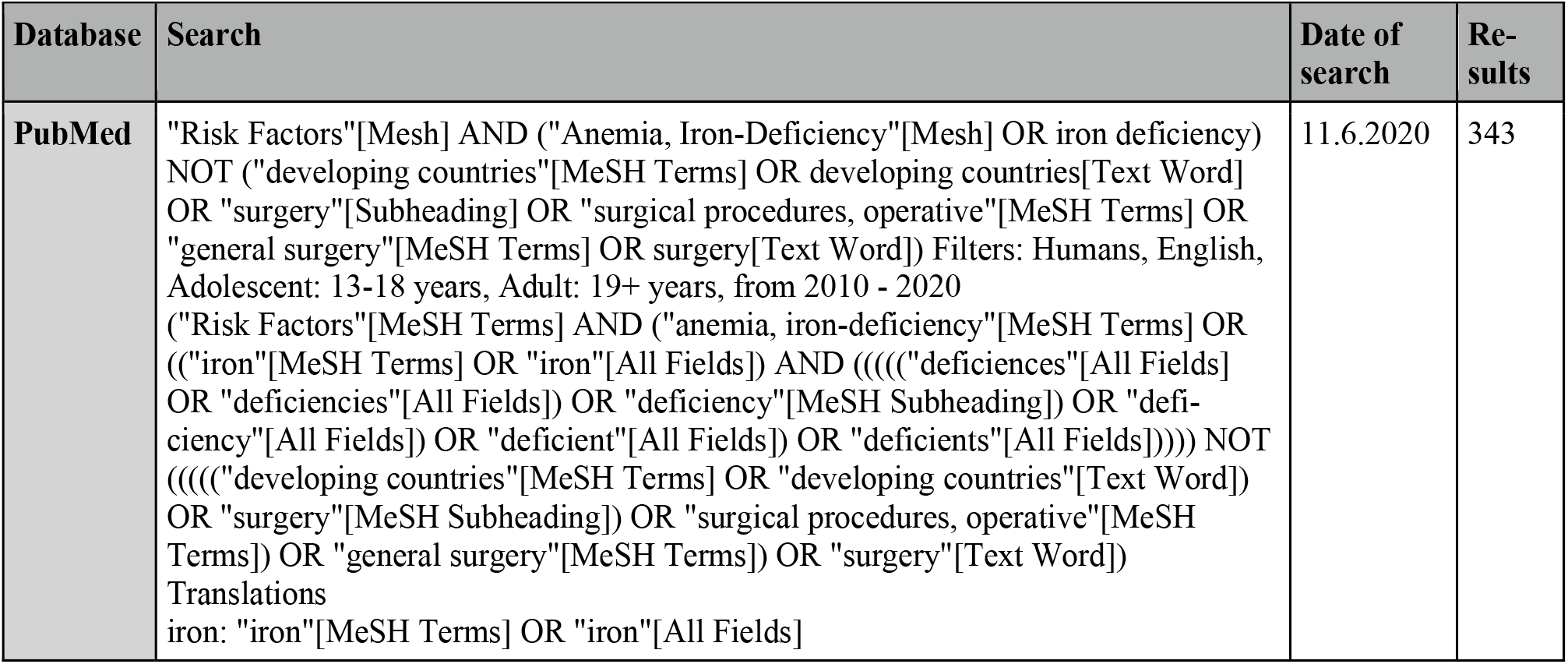
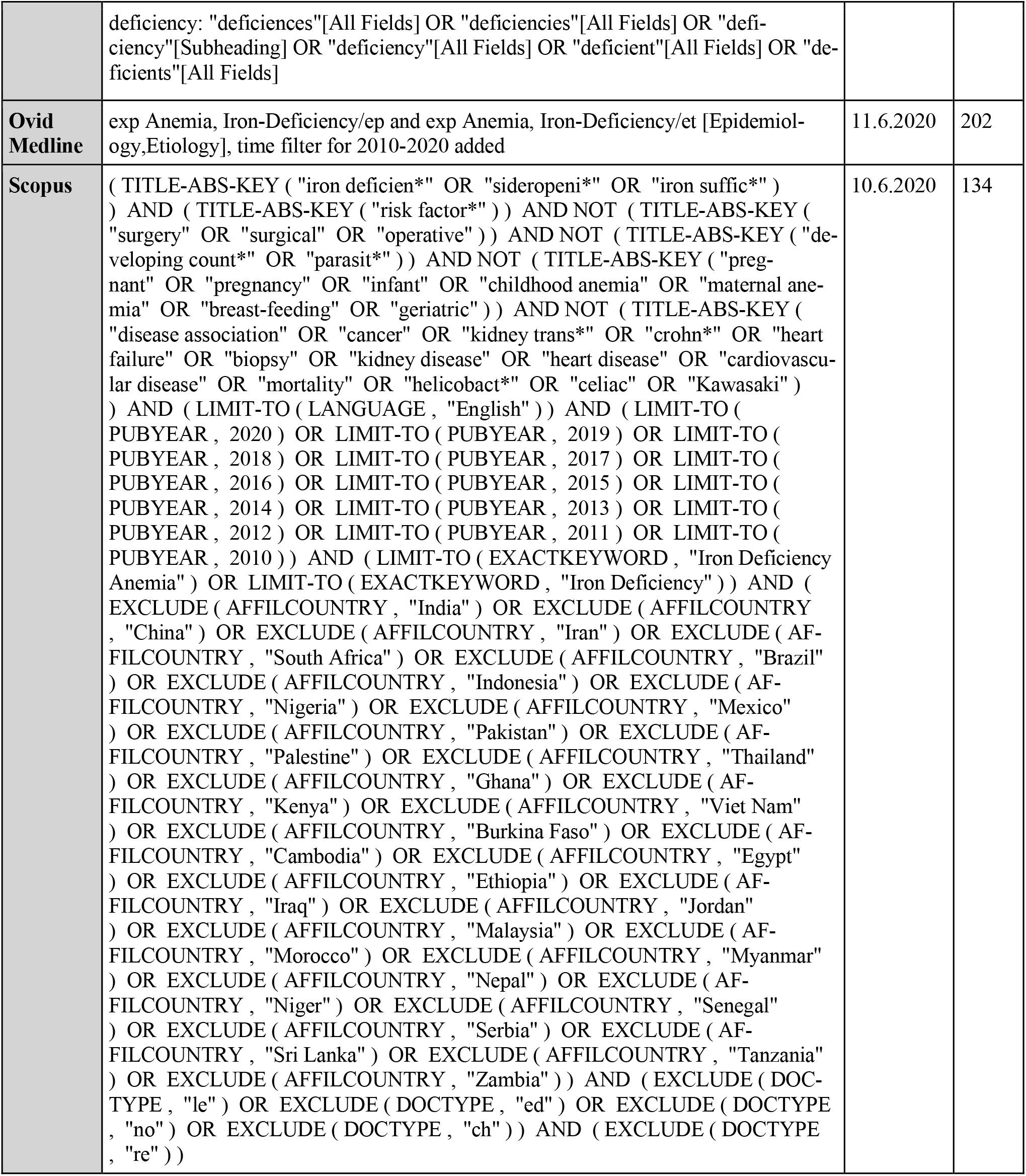
Search strategy

### Inclusion and exclusion criteria

As we were interested in current data from the past ten years, we decided to limit the search to articles published in English between the years 2010-2020. Some studies included participants younger than 15 years old, in these cases only data on the over 15-year-olds was considered. In order to produce a coherent data set we only included studies from countries with Human Development Index (HDI) ≥0.800 according to 2020 data (1).

We excluded articles that studied ID/IDA risk in patient populations with conditions that are considered impediments for blood donation, pregnancy, childbirth, surgical procedures and unlikely causes for ID/IDA in the European population (e.g. parasites). As we were interested in risk factors in a population of potential blood donors other than blood donation itself, we excluded studies concerning the effect of blood donation. We also excluded studies on genetic variants, as using them in blood donor selection processes currently is cumbersome.

Some articles study the etiology of other conditions, from these we derived rates of ID/IDA.

The search produced 679 abstracts in total. After screening based on the article title and abstract, articles were excluded as seen in Figure 1 and duplicates were removed. Full-text articles were then accessed and assessed for eligibility and articles were excluded as seen in Figure 1. The remaining 25 articles were included in analysis.

**Figure 1:**
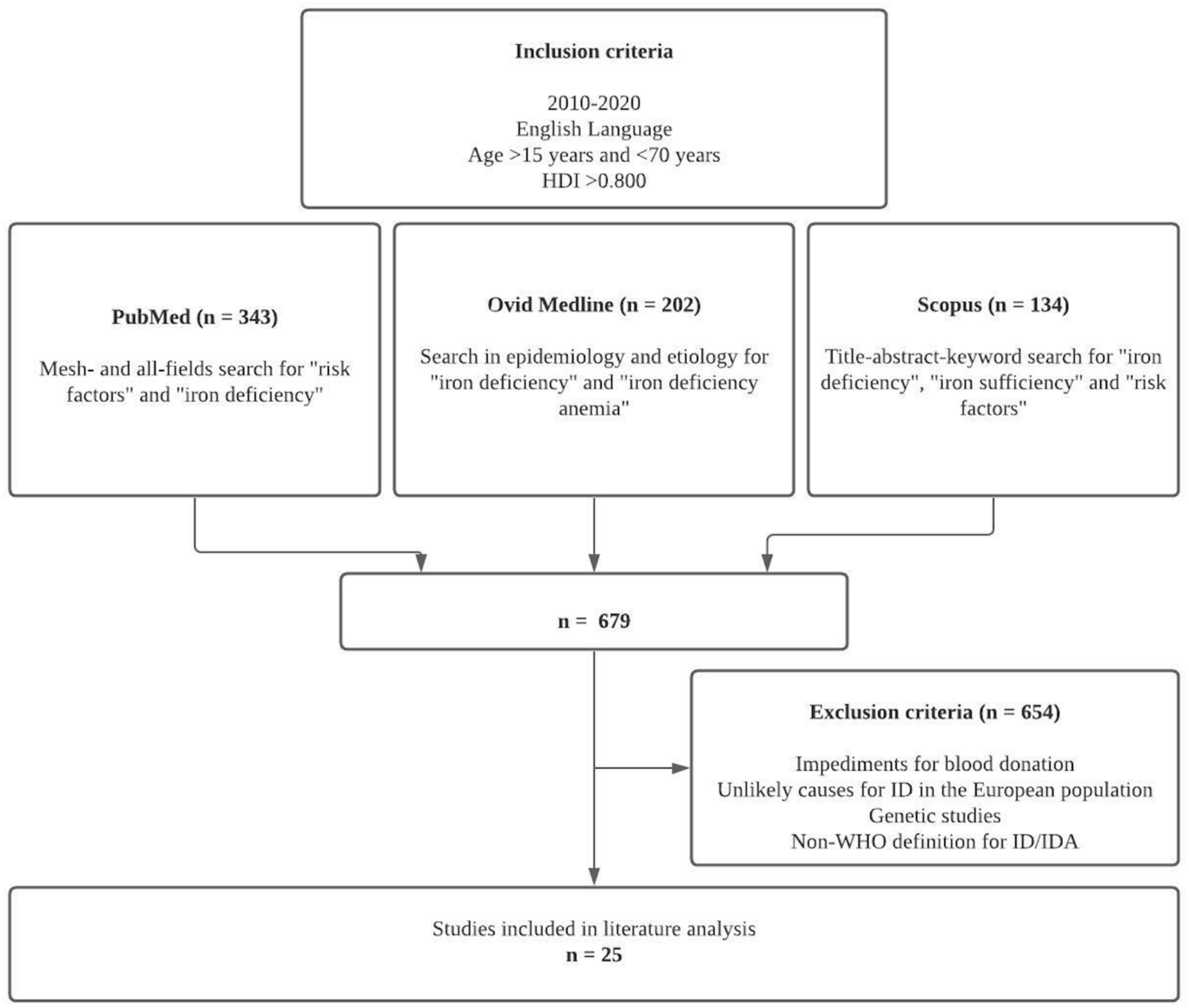

Iron deficiency was defined as serum ferritin ≤15 μg/l and iron deficiency anemia per the WHO definition of anemia of which most is caused by iron deficiency as hemoglobin ≤12 g/dl in women and ≤13 g/dl in men or appropriate ICD-9/ICD-10 codes (2).

## Results

A summary of the articles found in the literature search can be found in table 2. Studied variables that were not found to increase or decrease the risk of ID/IDA, or showed statistical difference but not in a statistically significant way are displayed in the “other factors studied” column. The column also includes variables that were investigated by the authors of the study, but we had excluded from our search.

**Table 2:**
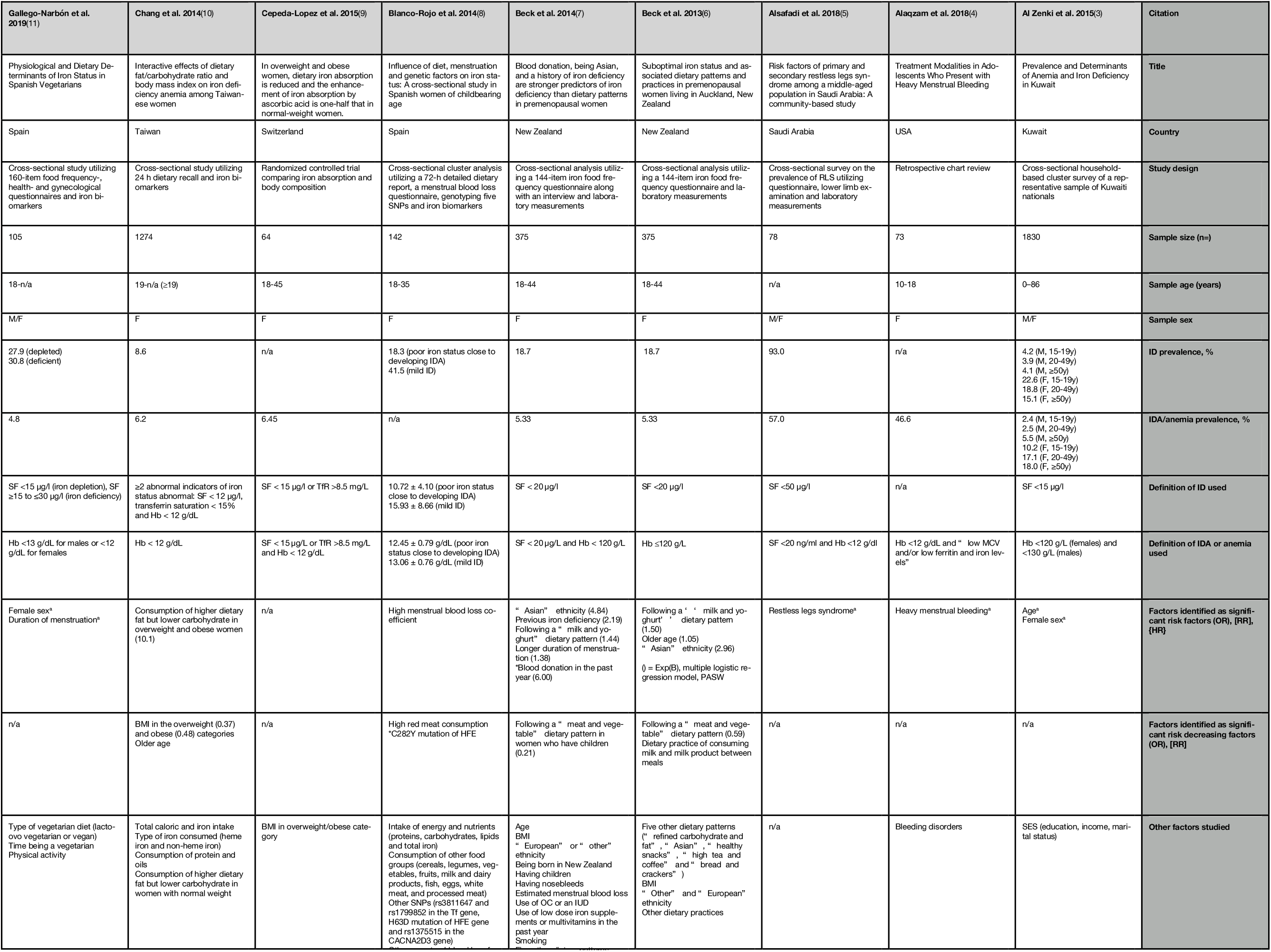
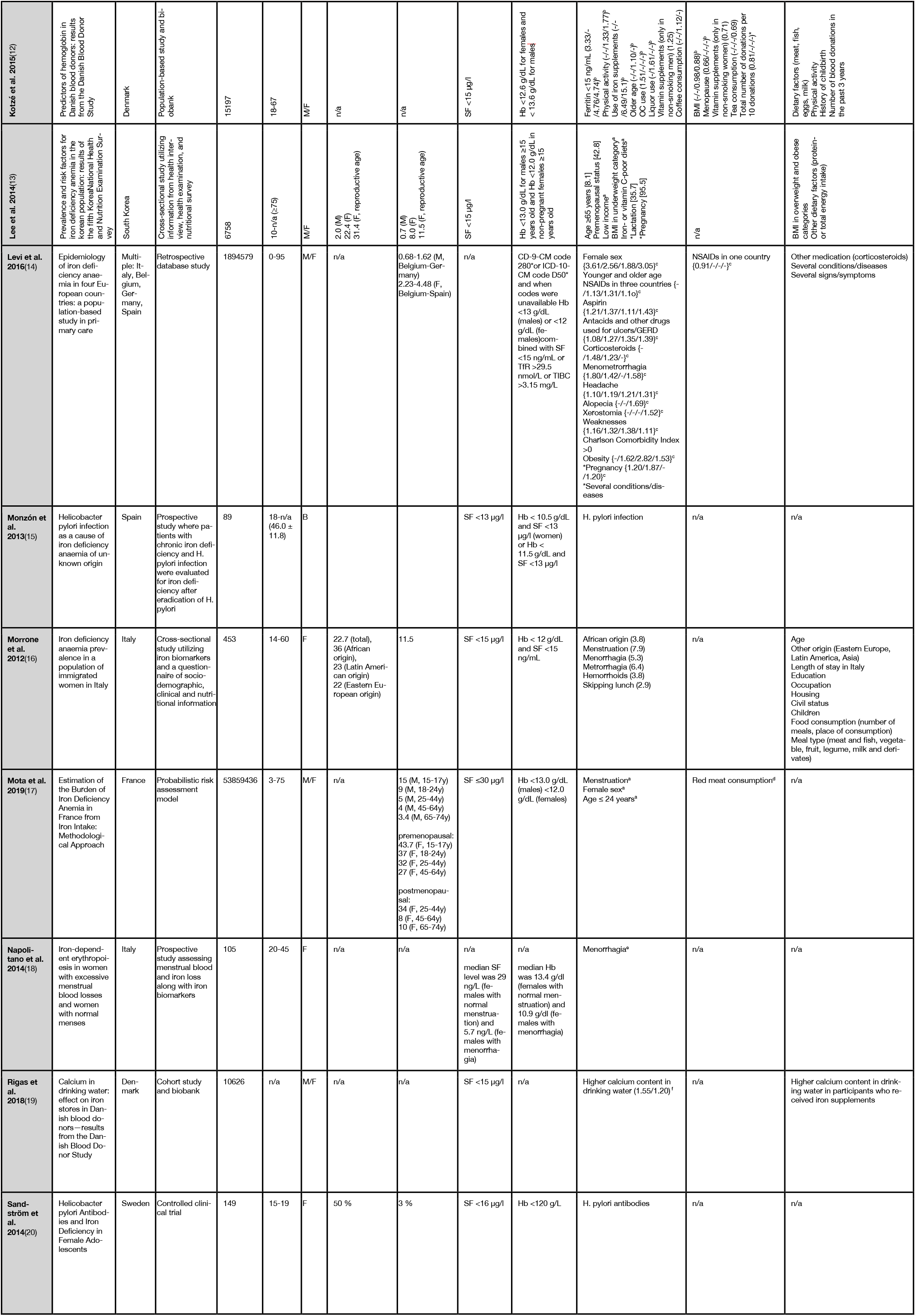
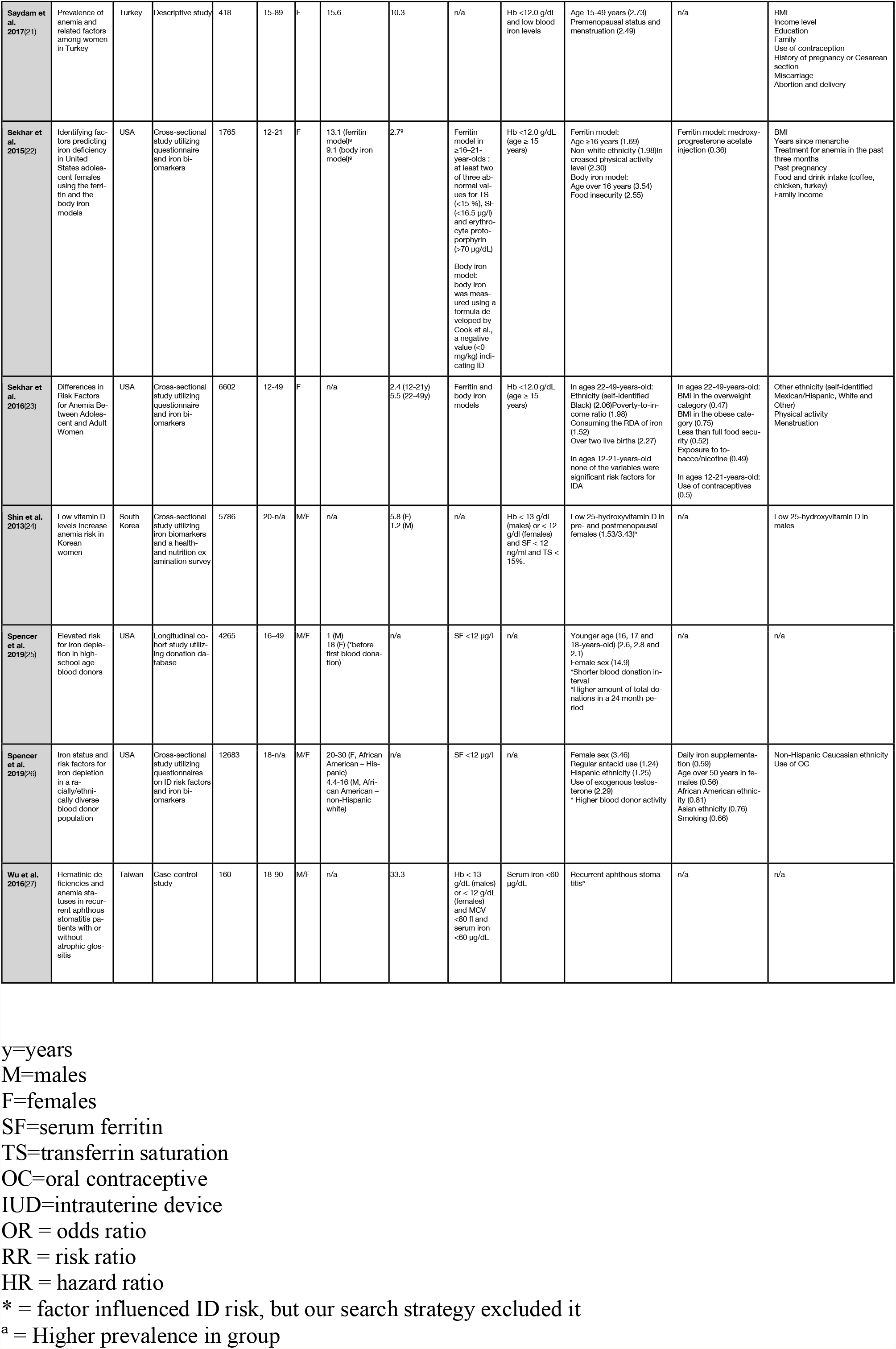
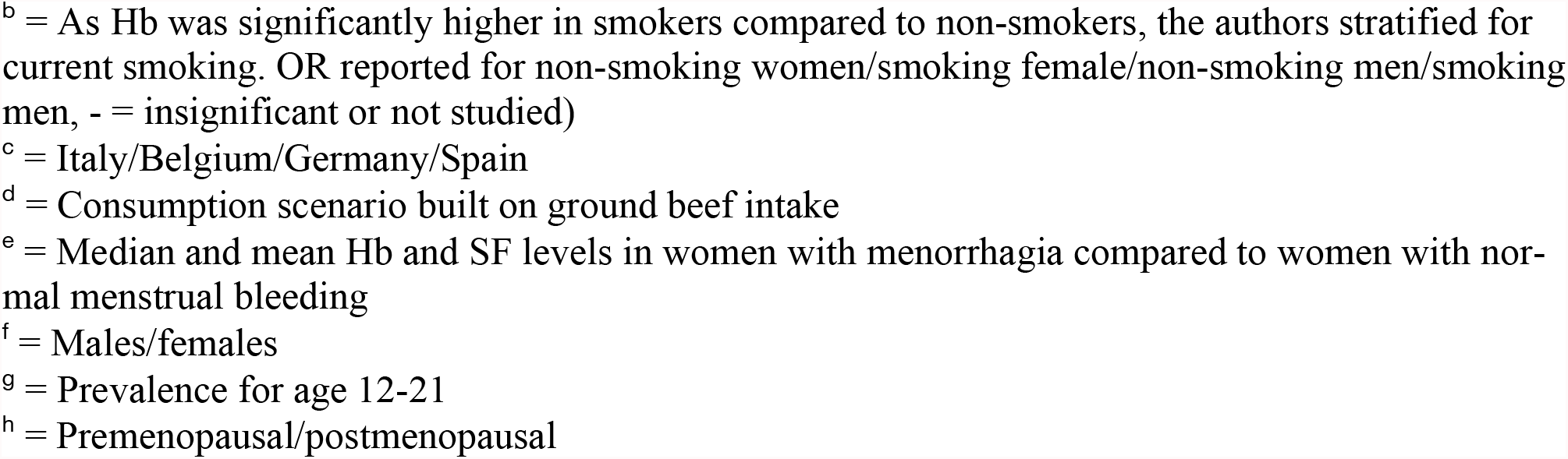
Study characteristics

### Sex

Overall, the prevalence of ID and IDA were significantly higher in females compared to males (3)(11)(13)(14)(17)(25)(26). One study concluded that there are, however, differences in the risk factors even among reproductive aged women by successfully identifying several significant risk factors in the older population (22-49 years) but not finding any variables for predicting IDA in the younger study population (12-21 years) (23).

### Menstruation

ID and IDA were found to be associated with menstrual blood loss, both when menstrual blood loss was self-reported and by measured blood loss(7)(8)(14)(16)(18)(21). Menstruation in women aged 25-44 years was the greatest contributor to the disease burden of IDA (DALY, disease adjusted life year) (17). The amount of years menstruating was also associated with IDA(23).

Heavy or prolonged menstrual bleeding (menorrhagia) was reported as a significant risk factor for ID and IDA in several studies (4)(7)(16) with evidence suggesting the IDA is caused by the heavy menstrual bleeding itself, not an underlying bleeding disorder (4). Evidence also showed a correlation between menstrual blood loss and menstrual iron loss, as well as a significantly higher iron loss in women with menorrhagia compared to normal menstruation. The study also found that most women with menorrhagia had depleted iron stores(18). Intermenstrual bleeding (metrorrhagia) also caused an increased risk of IDA (16). IDA risk was unsurprisingly also higher when both conditions were combined, as in menometrorrhagia (14).

As expected menopause and non-physiological amenorrhea decreased the risk for both ID and IDA(12)(13)(17).

### Age

In males ID prevalence remained similar across age groups showing a slight U-formed relationship, while IDA rates increased slightly with age (3)(13). One of the studies found ID prevalence to be the highest in the younger age group and decreasing with age(17). For females prevalence of both ID and IDA was significantly higher in reproductive age compared to older age groups (3)(13)(16)(17)(21)(22).

When prevalence was reported for both sexes combined, IDA prevalence was higher in young and middle-aged participants, decreasing in the oldest age group(14)(26). In a study on young blood donors age 16-18 years was found to be an independent determinant for ID in both males and females (25).

### BMI

BMI in the overweight and obese category had a protective effect on IDA in females(10)(23), although the effect was reduced with consumption of high dietary fat and low carbohydrate (10). Likewise, higher BMI decreased the risk of lower Hb levels(12). Low BMI was associated with higher IDA risk compared to those with BMI in the normal-weight or obese categories (13).

There was, however, some discrepancy in the results. Iron absorption and the absorption-enhancing effect of ascorbic acid were lower in overweight and obese women compared to normal-weight women (9). Obesity was also associated with IDA in one study (14), yet no statistically significant association between BMI and ID/IDA was found in two of the studies(9)(21).

### Dietary factors

Consumption of red meat was widely cited as a protective factor against ID(6)(7)(8). Interestingly, one study showed that total iron consumption was similar among groups with normal iron status, mild ID and ID, while the only significant difference in food intake was red meat consumption, suggesting heam iron is an important determinant for iron status(8). It was estimated that red meat consumption of 100 g/day would decrease although not eliminate the disease burden of IDA, as well as decrease prevalence of ID in French females(17). Although ID prevalence was high in women following a vegetarian diet, there was not a significant difference between the different types of vegetarian diet or time as a vegetarian. Prevalence of IDA in both sexes and ID in males were low (11).

In premenopausal women, following a “meat and vegetable” dietary pattern decreased the risk while following a “milk and yoghurt” dietary pattern increased the risk of ID(6)(7). In another study, “meat and vegetables” only decreased the odds in women who had children(7). The women were also more likely to have sufficient iron status if milk products were consumed between meals(6).

Once again, the evidence was somewhat inconsistent. Nondietary factors were stronger indicators of ID than dietary factors in premenopausal women(7). Two studies did not find any association between any dietary factors (including meat and milk) and IDA(12)(16), although skipping lunch did increase the risk(16). No association was detected between total energy or protein intake and IDA, although there was an association with lower than the recommend nutritional intake of iron and vitamin C (13). Vitamin D deficiency increased the risk of IDA in females but not in males (24).

### Medical conditions and symptoms

ID and IDA was associated with helicobacter pylori infection(15)(20). IDA prevalence was high in patients with restless legs syndrome (RLS), as IDA is a main risk factor for RLS (5). Symptoms associated with IDA were weakness, headache, xerostomia, alopecia and irritability. The same study also identified Charlson comorbidity index >0 as a risk factor (14). Additionally, IDA risk was increased with hemorrhoids (16) and aphthous stomatitis, especially when associated with atrophic glossitis (27).

### Use of exogenous hormones, medications and supplements

Although OC use was higher in females who had sufficient iron status than those who did not (7)(8), it was suggested that when OC use was considered alongside other determinants for ID or when menstrual bleeding was controlled for that there was no additional effect of OC use on ID risk(7). On the other hand, in one study OC use was associated with lower Hb levels (12), although this study did not consider menstruation as a variable. Another two studies found use of contraceptives to be protective against IDA (23) and ID (26). In addition, medroxyprogesterone acetate injection showed a protective effect against ID (22). As for exogenous hormone use in males, testosterone was associated with increased risk of ID (26).

Use of aspirin increased the risk of IDA risk, as did use of NSAIDs. However, the increased risk for NSAIDs only applied to three of four countries studied, whereas the risk decreased in one of the countries(26). Additionally, there was evidence for increased risk for both ID and IDA with use of antacids and/or PPIs (14)(26).

Iron supplement use was higher in groups with ID or IDA (11)(12), as well as associated with IDA in one study (23). Another study found daily iron supplementation to be protective against ID, but not if supplementation was less than daily (26).

### Socioeconomic factors

IDA was associated with low income (13) and low family poverty-to-income ratio (23). However, other studies did not find a significant association with SES (income(3)(21), education(3) or employment status (16)). Food insecurity in terms of worrying about running out of food increased the risk of ID in one study (22), although the effect was the opposite for IDA with less than full food security being protective against it in a subsequent study from the same team (23).

The IDA risk was higher in women with more than two live births as opposed to two or under (23).

### Ethnicity

Ethnicity was widely cited as a determinant for ID/IDA risk, although the results were indecisive.

“Asian” ethnicity was cited as a risk factor for ID when compared to “European”and“other”ethnicities(6)(7).“Non-white” was cited as a risk factor for ID(22) and “Black” for IDA(23). On the other hand, “African American” and “Asian” blood donors had lower risk for ID than “non-Hispanic Caucasian” donors, while “Hispanic” donors were of higher risk.

IDA prevalence was high in a study of immigrated women in Italy, in which women of African descent were the most likely to suffer from IDA (16).

### Environmental and lifestyle factors

Although ID was associated with exercise only in one study that also included children aged 12-14 years(22), iron-sufficient participants were more likely to exhibit higher amounts of exercise while participants with IDA were more likely to be sedentary(11). Contrarily, in other studies higher physical activity was more common in the group with lower Hb levels(12) or the effect of exercise didn’t reach statistical significance(23).

Smoking and exposure to tobacco or nicotine reduced the risk of ID(23)(26), and smokers had significantly higher mean Hb than non-smokers(12).

Although the effect size was minor, one of the studies found that the higher the calcium content in drinking water is, the higher the ID risk is (19).

## Discussion

We found evidence that strengthened previously reported results on reasons behind low hemoglobin deferrals in blood donors, like female sex, increasing age, low body weight and Hispanic or African ethnicities. Furthermore, we identified several other determinants for ID and IDA that potentially could be used for more individualized selection of blood donors. These included heavy menstrual bleeding and some medications and medical conditions, low SES as well as genetic variants and ethnicity. In this section, we will discuss these other determinants further.

### Menstruation

Menstruation has long been considered a risk factor for ID/IDA and it is evident from the studies in this review that heavy menstruation causes even higher rates of ID and IDA. It would be useful to further study the population of female donors to see if it is possible to tailor a more individualized approach for donor selection to minimize the risk in women who experience heavy menstrual bleeding.

Dietary factors are notoriously hard to study, which was highlighted in this review. Several possible risk factors for and protective factors against ID and IDA were identified in the studies included in this review, although none other than red meat and milk consumption stood out in terms of congruence in the multiple articles. While red meat consumption had a protective effect against ID and IDA, milk consumption increased the risk. Similarly, we have questioned participants on their dietary habits as part of a 28-question questionnaire in a prospective longitudinal cohort study on Finnish blood donors and found that the strongest association with higher ferritin levels was red meat consumption.

The effect of BMI and ethnicity were inconclusive due to contradictory results. Weight is commonly included in the donor selection process in order to offset low blood volume, but as far as we know BMI categories are not accounted for in donor selection processes. We have had some earlier evidence suggesting donors with higher BMI have higher ferritin levels in the FinDonor 10 000 study (29), but this review suggest there might be confounding factors not yet considered. Further study is also required to see the extent ethnicity causes variation in the blood donor population, preferably in combination with genetic studies. Blood donors from all ethnic backgrounds are needed due to rare blood groups, and it is vital that the donor population is varied.

Several medications were associated with lowered iron parameters, significantly PPIs and antacids. Gastric acid facilitates absorption of iron and ID has been linked to PPI use in the past (30). Therefore, asking donors about use of either of these types of medications could be warranted, although further inquiry into the magnitude of the dose-effect relationship is and how fast iron storage recovers after the medication is discontinued is needed. As far as having a protective effect use of hormonal contraception did show an association for women with heavy menstrual bleeding and could possibly also be used as a guiding factor in the selection process, possibly in relation to heavy menstruation discussed earlier. NSAIDs and aspirin use were also identified as risk factors, with aspirin showing the same risk in all four countries studies while results for NSAIDs were more inconclusive. One explanation for this result could be the secondary preventative use of low-dose aspirin and possible concurrent use of PPIs and NSAIDs.

Although lower SES was a risk factor for ID and IDA, comparison between different countries is difficult due to different social and economic settings and using SES as a factor for donor selection is impractical. Vitamin D deficiency and water hardness do not seem to be a significant enough cause to warrant change in the blood donor selection process. RLS shows some potential as a marker for IDA, although further research is needed to assess if symptoms resolve or persist after successful IDA treatment.

### Study strengths and limitations

There are several methodological limitations to this study.

Firstly, the studies used were not limited to just one measure for ID or IDA. The iron status was assessed by ferritin and Hb concentrations and ICD codes. Some studies used CRP to confirm non-inflammatory status, but this was not the case in all studies. Therefore, it is difficult to determine how the results relate to one another. Secondly, the studies used different definitions for ID. Thirdly, the study designs in the records used differed from each other. This is not necessarily a bad thing as it gives us a larger perspective of the studies done on this subject. Nonetheless, it makes comparison inconvenient.

One major methodological flaw of this study is that H. Pylori infection was excluded from the Scopus search in the early stages in order to get a more manageable search result. This likely excluded some potential articles on these subjects. However, these terms were not excluded from the PubMed and Ovid Medline searches.

Possible overlapping of risk factors is a challenge that this study was not able to answer. These confounding factors have not been accounted for in this review and further investigation is needed to determine the actual prevalence of ID and IDA when controlling for these factors. The results can also be skewed due to the so-called healthy donor effect (31).

The strengths of this study are the broad use of search terms as well as the use of three different databases.

In conclusion, current evidence suggests that several determinants, including biological, environmental, lifestyle and socioeconomic factors increase the risk for ID. Although several of these factors are commonly accounted for in blood donor selection, we identified some determinants that show potential for inclusion in the selection process. These include heavy menstrual bleeding and use of some medications. Further study is needed to ascertain whether inclusion would influence donor health and the blood supply as well as how best to implement the changes if they are deemed necessary.

## Data Availability

All data can be reproduced by following the search guidelines

